# The reactogenicity and immunogenicity of a booster dose after the second dose of a protein subunit vaccine MVC-COV1901: An extension to an open-label, dose-escalation, and non-randomized phase 1 study

**DOI:** 10.1101/2021.12.01.21267115

**Authors:** Szu-Min Hsieh, Shan-chwen Chang, Hao-Yuan Cheng, Shin-Ru Shih, Chia En Lien

## Abstract

The waning antibody levels after immunization and the emergence of SARS-CoV-2 variants of concern (VoC) negatively impact the vaccine-induced neutralization of SARS-CoV-2. In this extension to the phase 1 clinical study, we report the antibody durability until 180 days after the second dose of MVC-COV1901, and examined the reactogenicity and immunogenicity followed by a booster shot MVC-COV1901 administered to 45 healthy adults from 20 to 49 years of age on day 209. Adverse reactions after the booster dose were mostly mild and comparable to that of the first two doses. Neutralizing antibodies remained detectable on day 209 at 59.4, 79.4, and 113.2 (IU/mL) for low dose (LD), middle dose (MD), and high dose (HD) groups, respectively. At four weeks after the booster dose, neutralizing titers increased to 1719.6, 818.3, and 1345.6 for LD, MD, and HD groups, respectively. Our data also showed that three doses of MVC-COV1901-induced antibodies were still effective, albeit lowered neutralizing titers, against the Omicron variant.

## Introduction

The use of booster vaccines in fully vaccinated individuals have been implemented around the world in response to surging cases due to waning immunity and infection by variants of concern (VoCs) such as Delta and Omicron [1, 2]. Boosting with same type of vaccine (homologous) or different type of vaccine (heterologous) to the primary series of vaccination has shown to be effective in enhance immunogenicity against VoCs [3-5]. MVC-COV1901 is a protein subunit COVID-19 vaccine based on the stable prefusion spike protein S-2P adjuvanted with CpG 1018 and aluminum hydroxide that has been authorized for emergency use in Taiwan [6]. The phase 1 clinical trial of MVC-COV1901 was designed as a dose-escalation trial with three groups of dose levels at 15 subjects for each group [7]. The current study is an extension of the MVC-COV1901 phase 1 study to investigate the antibody persistence after the second dose and the immunogenicity and reactogenicity of a booster dose six months after primary vaccination.

## Methods

This is an extension to the phase 1 study to evaluate the safety and immunogenicity of MVC-COV1901 [7]. Three different dose levels employed in the original Phase 1 trial were low dose (LD), middle dose (MD), and high dose (HD) (5 μg, 15 μg, and 25 μg, respectively) of S-2P protein adjuvanted with CpG 1018 and aluminum hydroxide [7]. Booster doses were administered on Day 209 with MD used as the booster dose for the LD and MD group, while HD was used for the HD group (Figure 1).

**Figure 1.**
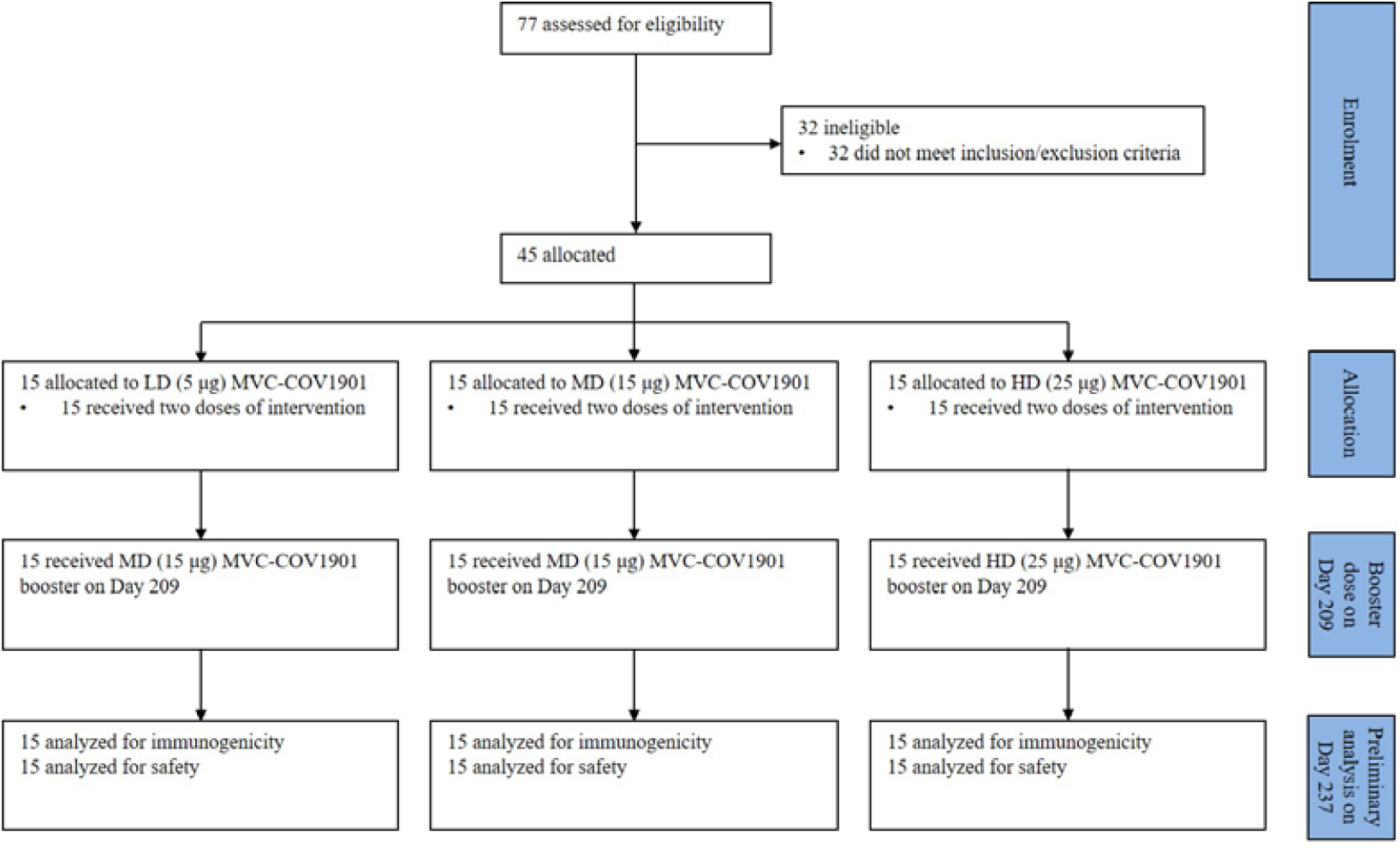
CONSORT flow diagram for the study.

Anti-SARS-CoV-2 spike antibody levels were measured by enzyme-linked immunosorbent assay (ELISA) using customized 96-well plates coated with S-2P antigen and expressioned in Binding Antibody Unit per mL (BAU/mL) [7]. Neutralizing antibody levels were measured by live-SARS-CoV-2 (Wuhan wildtype, hCoV-19/Taiwan/4/2020, GISAID# EPI_ISL_411927) neutralization assay previously performed [6]. Briefly, two-fold serial dilutions of serum samples were mixed with equal volume of 100 TCID_50_/50 μL of virus and incubated at 37 °C for one hour. After incubation, the mixture was added to Vero E6 cells and incubated at 37 °C for four to five days. The results were expressed in WHO Standardized International Unit per mL (IU/mL, WHO Standardized) according to previously established methods [6]

The serum samples from MD and HD groups were tested in pseudovirus neutralization assay against pseudoviruses with spike protein of Wuhan wildtype and Omicron variant constructed and performed as in the phase 1 clinical study [7]. Two-fold serial dilution of serum samples were mixed with equal volume of pseudovirus and incubated before adding to the HEK-293-hAce2 cells. Fifty percent inhibition dilution titers (ID_50_) were calculated with uninfected cells as 100% neutralization and cells transduced with virus as 0% neutralization. The mutations for Omicron variant used in the spike sequence for pseudovirus construction are: A67V, del69-70, T95I, G142D, del143-145, del211, L212I, ins214EPE, G339D, S371L, S373P, S375F, S477N, T478K, E484A. Q493R, G496S, Q498R, N501Y, Y505H, T547K, D614G, H655Y, N679K, P681H, D796Y, N856K, Q954H, N969K, L981F according to the most common combination of mutations listed on Coronavirus3D (https://coronavirus3d.org/)

## Results

Solicited adverse effects (AEs) are shown as in Figure 2. No SAE or AESI were reported and reactogenicity after the third dose for all participants was comparable to that after the previous two doses. The most common solicited local and systemic effects after any dose were pain and malaise/fatigue, respectively. In terms of solicited local AEs, booster dose resulted in slightly more elevated incidences of pain compared to the first dose and the second dose. A notable difference in solicited systemic AE is the incidence of two events of fever each of grade 1 and 2 in the MD and HD groups after the booster dose, whereas no incidences of fever were reported in the first two doses.

**Figure 2.**
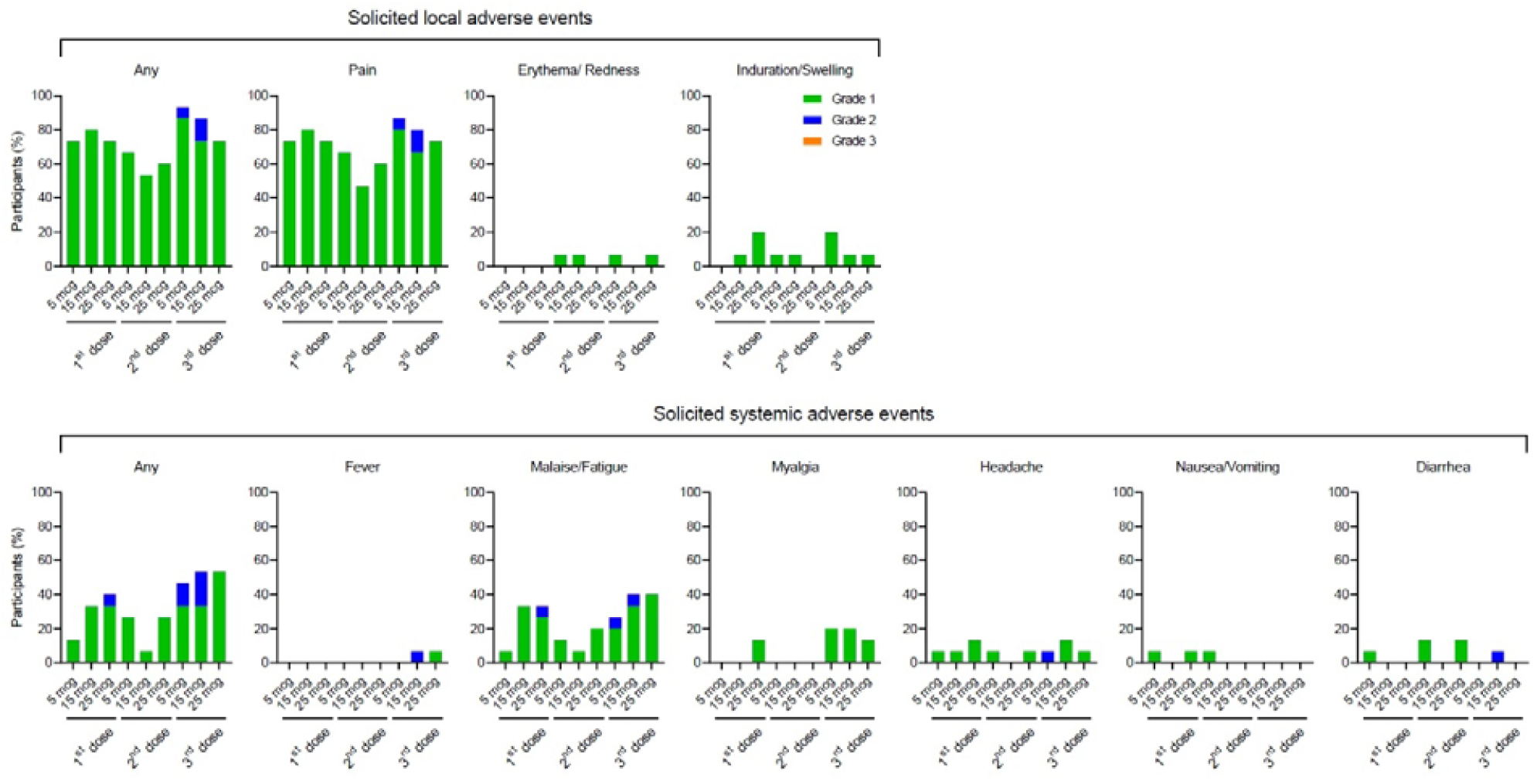
Solicited local and systemic adverse events after 1st, 2nd, and 3rd dose of MVC-COV1901

Anti-SARS-CoV-2 antibody levels persisted in all groups at day 209 (6 months after the second vaccination), with geometric mean titers (GMTs, in BAU/mL) of 40.1, 59.9, and 86.6 for LD, MD, and HD groups, respectively (Figure 3A and Appendix A). The corresponding neutralizing NT_50_ GMTs (IU/mL) were 59.4, 79.4, and 113.2 for LD, MD, and HD groups, respectively (Figure 3B and Appendix B). At four weeks after the booster dose (day 237), the neutralizing GMTs (IU/mL) of the live-virus SARS-CoV-2 neutralizing assay were 1719.6, 818.3, and 1345.6 for LD, MD, and HD groups, respectively (Figure 3B and Appendix B). Serums samples from day 237 of MD and HD groups were subjected to wildtype and Omicron variant pseudovirus assay. Compared to the wildtype, Omicron variant resulted in 8.7- and 6.4-fold decrease in neutralizing antibody titer levels for the MD and HD groups, respectively (Figure 4). All of individual titer levels in the HD group were above the lower limit of detection, while one individual in the MD group had undetectable neutralizing titer against the Omicron variant (Figure 4).

**Figure 3.**
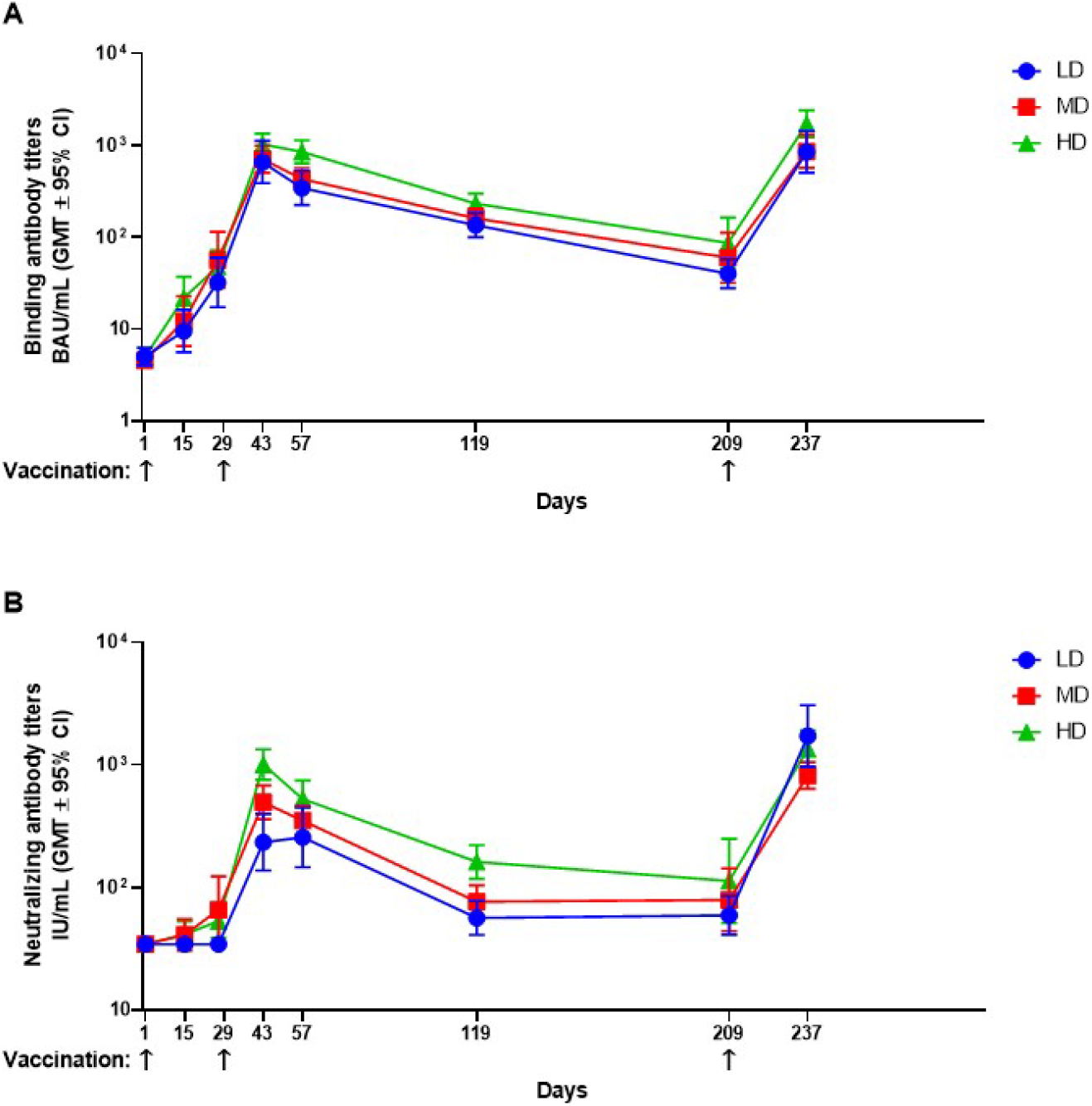
Immunogenicity of MVC-COV1901 during the study course. Arrows at the bottom indicate the time of vaccination. Results are expressed as symbols representing the GMT and error bars representing the 95% confidence interval of **A)** anti-SARS-CoV-2 IgG antibody titer expressed in binding antibody titers (BAU/mL) and **B)** neutralizing antibody titer expressed in WHO International Units (IU/mL) of LD, MD, and HD groups at various time points.

**Figure 4.**
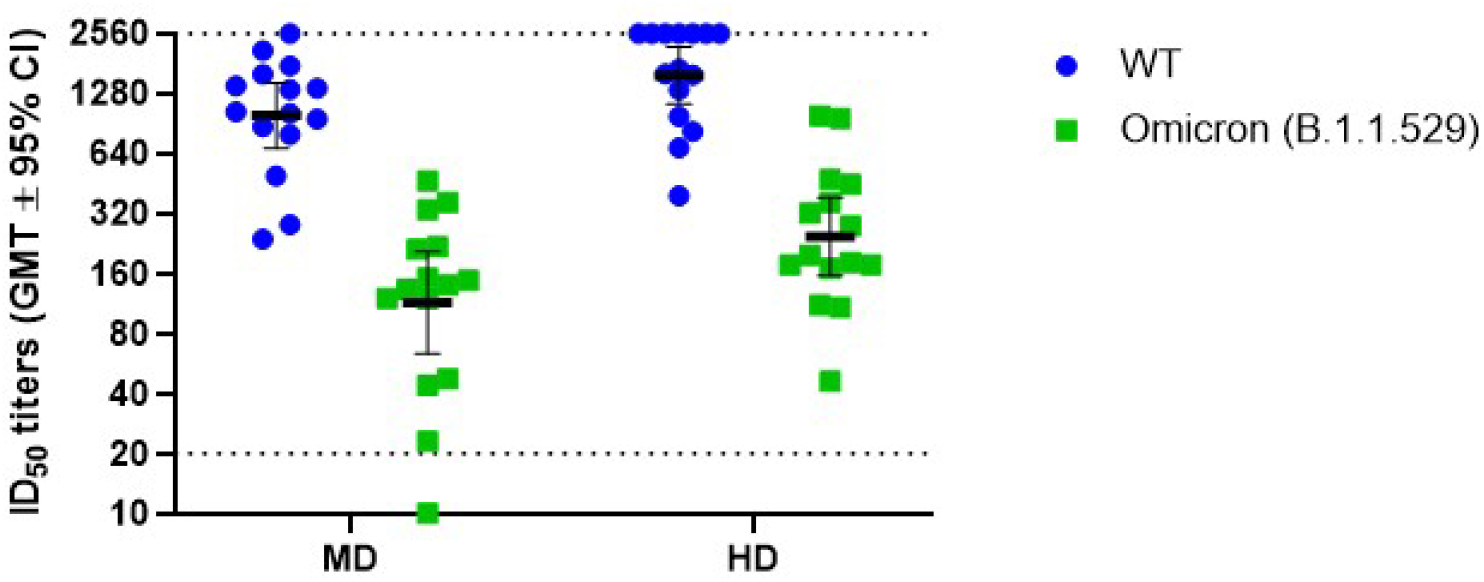
Pseudovirus neutralization assay of pseudovirus with spike proteins of Wuhan wildtype (WT) or Omicron variant with serum samples from the MD and HD groups. Results are presented as symbols representing the individual ID_50_ titers and the black horizontal bar representing the ID_50_ GMT with error bars representing the 95% confidence interval. Dotted lines indicate upper and lower limits of detection.

## Discussion

Although the Correlates of Protection (CoP) is yet to be established for COVID-19 vaccines, neutralizing antibody titers have been reported to be highly correlated with vaccine efficacy [8]. Based on Khoury et al’s modelling, a level of neutralizing NT_50_ 54 IU/mL could render 50% of vaccine efficacy against the prototype strain, which meets the WHO target product profile for COVID-19 vaccine approval. The trough, at 6 months after the primary two doses of MVC-COV1901, of neutralizing NT_50_ for MD group was 79.4 IU/mL, which still exceed the above cut-off. The dynamics of antibody titer after the second dose is similar to that of other vaccines, namely increasing to peak at 2 weeks after the administration and decline afterwards but remain detectable on day 209 [9]. The GMT of neutralizing antibody titers of MD showed a 6.2 fold-reduction (495.9/79.4) at 180 days - which is comparable to other platforms - compared to the peak at 14 days after the second dose [10, 11]. At four weeks after the booster dose, a fold-increase of 1.7 (818.3/495.9) was noted compared to the previous peak. The results showed that antibody response against SARS-CoV-2 can be boosted by the third dose.

We have examined the immunogenicity of antibodies induced by three doses of MVC-COV1901 against the emerging Omicron variant. As in Figure 4, the neutralizing antibody GMT against the Omicron variant after three doses of MVC-COV1901 was lower than that of the wildtype, although at the high dose level the fold reduction was 6.4, lower than that of the MD group (8.7-fold). This result is in line with recent data that Omicron variant is resilient against majority of anti-SARS-CoV-2 monoclonal antibody drugs and convalescent sera, and two doses of currently available vaccines performed poorly against the Omicron variant [12-15]. Administration of booster doses were able to increase neutralizing activities against the Omicron variant as evident by booster doses of recombinant protein subunit vaccine NVX-CoV2373 and Moderna/Pfizer mRNA vaccines compared to only two doses of vaccines [3, 14, 15]. The data here supports the use of the vaccine in further clinical development that involves a third dose boost as protection against the dominant Delta and Omicron variants. Ongoing monitoring is planned and will give insights to antibody persistence after the third dose.

## Data Availability

All data produced in the present study are available upon reasonable request to the authors.

## Conflict of Interest Disclosure

H.-Y.C. and C.E.L. are employees of Medigen Vaccine Biologics Corporation and have received grants from the Taiwan Centers of Disease Control, Ministry of Health and Welfare. S.-M.H., S.-C.C., and S.-R.S. declared no conflict of interest.

All authors have reviewed and approved of the final version of the manuscript.

## Author Contributions

Concept and design: S.-M.H.

Acquisition and interpretation of data: S.-M.H. and S.-C.C.

Drafting of the manuscript: S.-M.H., H.-Y.C., C.E.L., and S.-C.C.

Laboratory assays and data analysis: S.-R.S.

## Additional Contributions

The Institute of Biomedical Sciences, Academia Sinica performed the neutralization assay. Members of Medigen Vaccine Biologics Corp. assisted in manuscript editing and revision.

## Ethics Statement

The trial protocol and informed consent form were approved by the Taiwan Food and Drug Administration and the Research Ethics Committee of National Taiwan University Hospital. The trial was conducted in accordance with the principles of the Declaration of Helsinki and Good Clinical Practice. An independent data and safety monitoring board (DSMB) was established to monitor safety data and the trial conduct. This trial is registered at ClinicalTrials.gov: NCT04487210

