## Appendices A and B for "The reactogenicity and immunogenicity of a booster dose after the second dose of a protein subunit vaccine MVC-COV1901: An extension to an open-label, dose-escalation, and non-randomized phase 1 study" (from another version of this preprint: version 1, 2021-12-05)

**Appendix A. Binding Antibody Geometric Mean Titer (BAU/mL)**

| **Binding Antibody Geometric Mean Titer (BAU/mL)** | | | |
| --- | --- | --- | --- |
| **Geometric Mean Titer by Visit** | **5 mcg** | **15 mcg** | **25 mcg** |
| **Visit 2 (Baseline)** | | | |
| N (Missing) | 15 (0) | 15 (0) | 15 (0) |
| GMT (95% Confidence Interval) | 5.05  (4.06 ~ 6.29) | 4.56  (4.56 ~ 4.56) | 4.97  (4.13 ~ 5.99) |
| **Visit 4 (Day 15)** | | | |
| N (Missing) | 15 (0) | 15 (0) | 15 (0) |
| GMT (95% Confidence Interval) | 9.55  (5.61 ~ 16.24) | 12.25  (6.57 ~ 22.83) | 21.94  (12.96 ~ 37.14) |
| **Visit 5 (Day 29)** | | | |
| N (Missing) | 15 (0) | 15 (0) | 15 (0) |
| GMT (95% Confidence Interval) | 32.24  (17.35 ~ 59.91) | 56.76  (28.14 ~ 114.48) | 47.93  (31.82 ~ 72.18) |
| **Visit 7 (Day 43)** | | | |
| N (Missing) | 15 (0) | 15 (0) | 15 (0) |
| GMT (95% Confidence Interval) | 654.66  (386.72 ~ 1108.23) | 706.44  (504.36 ~ 989.50 | 1023.31  (783.62 ~ 1336.34 |
| **Visit 8 (Day 57)** | | | |
| N (Missing) | 15 (0) | 15 (0) | 15 (0) |
| GMT (95% Confidence Interval) | 343.22  (224.05 ~ 525.80) | 430.39  (328.36 ~ 564.13) | 848.03  (637.37 ~ 1128.28) |
| **Visit 9 (Day 119)** | | | |
| N (Missing) | 15 (0) | 15 (0) | 15 (0) |
| GMT (95% Confidence Interval) | 135.06  (99.90 ~ 182.60) | 160.38  (126.12 ~ 204.00) | 231.33  (179.12 ~ 298.76) |
| **Visit 10 (Day 209)** | | | |
| N (Missing) | 15 (0) | 15 (0) | 15 (0) |
| GMT (95% Confidence Interval) | 40.13  (27.89 ~ 57.74) | 59.85  (32.02 ~ 111.88) | 86.60  (45.93 ~ 163.29) |
| **Visit 12 (Day 237)** | | | |
| N (Missing) | 15 (0) | 15 (0) | 15 (0) |
| GMT (95% Confidence Interval) | 843.61  (502.94 ~ 1415.01) | 853.94  (568.31 ~ 1283.11) | 1704.82  (1216.81 ~ 2388.55) |

**Appendix B. Neutralizing Antibody Geometric Mean Titer (IU/mL)**

| **Neutralizing Antibody Geometric Mean Titer (IU/mL)** | | | |
| --- | --- | --- | --- |
| **Geometric Mean Titer by Visit** | **5 mcg** | **15 mcg** | **25 mcg** |
| **Visit 2 (Baseline)** | | | |
| N (Missing) | 15 (0) | 15 (0) | 15 (0) |
| GMT (95% Confidence Interval) | 34.47  (34.47 ~ 34.47) | 34.47  (34.47 ~ 34.47) | 34.47  (34.47 ~ 34.47) |
| **Visit 4 (Day 15)** | | | |
| N (Missing) | 15 (0) | 15 (0) | 15 (0) |
| GMT (95% Confidence Interval) | 34.47  (34.47 ~ 34.47) | 41.10  (30.59 ~ 55.21) | 41.57  (32.62 ~ 52.96) |
| **Visit 5 (Day 29)** | | | |
| N (Missing) | 15 (0) | 15 (0) | 15 (0) |
| GMT (95% Confidence Interval) | 34.47  (34.47 ~ 34.47) | 66.09  (35.29 ~ 123.75) | 53.39  (38.83 ~ 73.41) |
| **Visit 7 (Day 43)** | | | |
| N (Missing) | 15 (0) | 15 (0) | 15 (0) |
| GMT (95% Confidence Interval) | 234.03  (137.69 ~ 397.78) | 494.85  (360.54 ~ 679.19) | 1006.42  (756.45 ~ 1338.99) |
| **Visit 8 (Day 57)** | | | |
| N (Missing) | 15 (0) | 15 (0) | 15 (0) |
| GMT (95% Confidence Interval) | 256.20  (146.64 ~ 447.61) | 351.17  (263.21 ~ 468.51) | 527.56  (373.03 ~ 746.13) |
| **Visit 9 (Day 119)** | | | |
| N (Missing) | 15 (0) | 15 (0) | 15 (0) |
| GMT (95% Confidence Interval) | 56.46  (41.13 ~ 77.51) | 76.59  (56.42 ~ 103.97) | 161.82  (117.92 ~ 222.05) |
| **Visit 10 (Day 209)** | | | |
| N (Missing) | 15 (0) | 15 (0) | 15 (0) |
| GMT (95% Confidence Interval) | 59.40  (41.24 ~ 85.54) | 79.35  (44.04 ~ 142.97) | 113.16  (51.35 ~ 249.34) |
| **Visit 12 (Day 237)** | | | |
| N (Missing) | 15 (0) | 15 (0) | 15 (0) |
| GMT (95% Confidence Interval) | 1719.56  (965.79 ~ 3061.63) | 818.31  (637.37 ~ 1050.61) | 1345.55  (946.81 ~ 1912.21) |
