## Supplementary figures and images for "The reactogenicity and immunogenicity of a booster dose after the second dose of a protein subunit vaccine MVC-COV1901: An extension to an open-label, dose-escalation, and non-randomized phase 1 study" (from another version of this preprint: version 1, 2021-12-05)

### Appendix C

**Appendix C. Solicited local and systemic adverse events after 1^st^, 2^nd^, and 3^rd^ dose of MVC-COV1901**

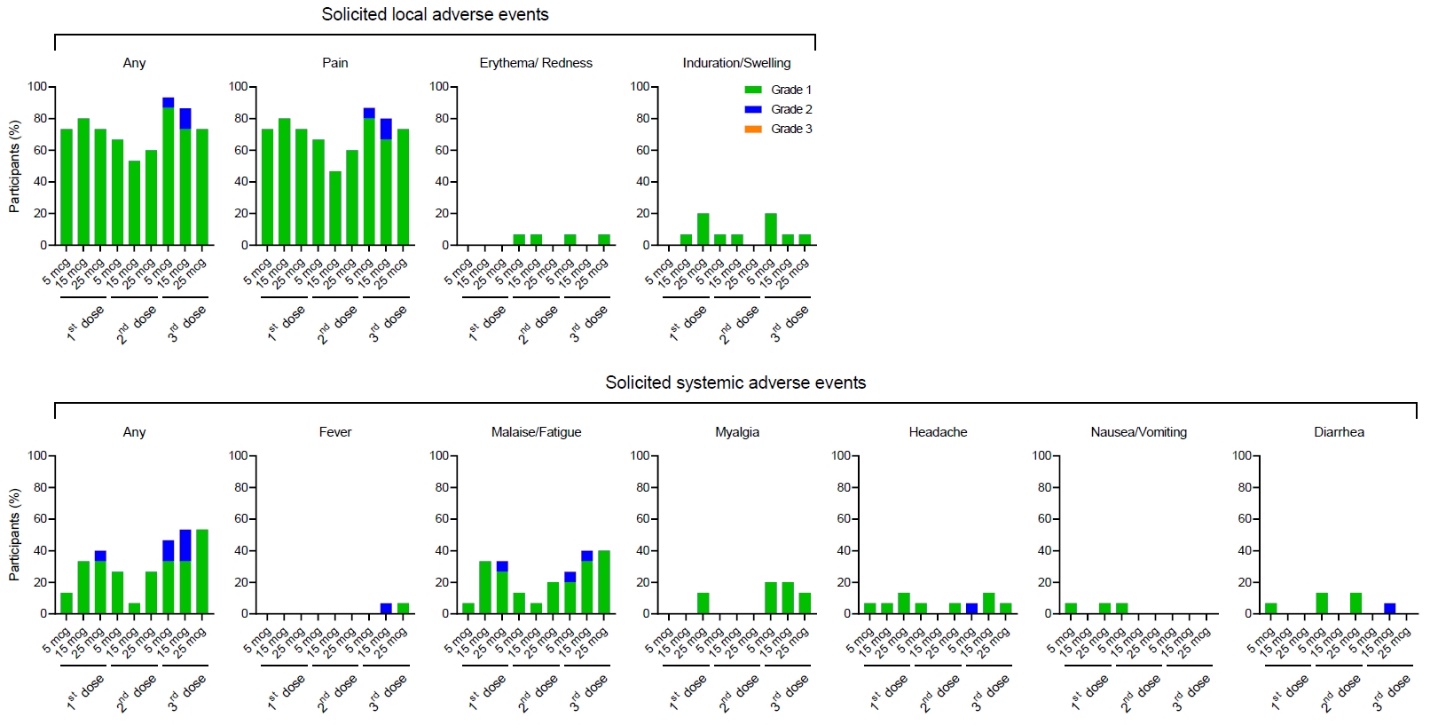
